# Lenvatinib plus pembrolizumab for the Treatment of Patients With Non-BRAF mutated Anaplastic Thyroid Cancer: A single center, phase 2 clinical trial

**DOI:** 10.1101/2025.11.09.25339786

**Authors:** Maria E. Cabanillas, Naifa L. Busaidy, Mark Zafereo, Priyanka C. Iyer, Jennifer R. Wang, Sarah Hamidi, Renata Ferrarotto, Suyu Liu, Gary B. Gunn, Michael Spiotto, Anastasios Maniakas, Maria Gule-Monroe, Victoria E. Banuchi, Mimi I. Hu, Matthew Ning, Ramona Dadu

## Abstract

**Background:** Dabrafenib (BRAF inhibitor) plus trametinib (MEK inhibitor) are approved for BRAF V600E-mutated anaplastic thyroid cancer (ATC), but ∼60% of tumors do not harbor BRAF V600E, leaving these patients without effective treatment options. The combination of lenvatinib + pembrolizumab was studied in the ATLEP trial in Europe, demonstrating a response rate of 52%, and median progression-free survival (PFS) and overall survival (OS) of 10 and 11 months, respectively, in ATC.

**Methods:** This was a phase 2 study of patients with BRAF wild type ATCs were treated with concurrent lenvatinib 20 mg po daily and pembrolizumab 400 mg IV Q6 weeks. Those at high risk of bleeding could start on a reduced dose of lenvatinib. The primary endpoint was median OS and secondary endpoints were response rate and PFS. With historical median OS of 3 months with single agent lenvatinib, the trial aimed to improve OS by additional 3 months.

**Results:** Twenty-five patients with a median age of 62 years were enrolled, of which 64% were men. All patients had distant metastases at study entry. With a median follow-up time of 17.5 months (range 8.8-41.4) for alive patients, the median OS and PFS were 11.4 (95%CI: 7.8 to 35.6; p<0.025), and 5.4 months (95%CI: 3.8-11.0), respectively. Best overall response in target lesions was 36%, including 1 complete and 8 partial responses.

**Conclusion:** Lenvatinib + pembrolizumab demonstrates efficacy in patients with metastatic, non-BRAF mutated ATC and should be considered a standard of care in this patient population.

## Introduction

Anaplastic thyroid cancer (ATC) is a rare and rapidly fatal form of thyroid cancer. Survival in these patients is driven primarily by stage at presentation and the BRAFV600E mutation status^1^. The most effective treatment for BRAFV600E mutated ATC is the BRAF/MEK inhibitor combination, dabrafenib/trametinib^2^ (FDA approved in 2018) +/-pembrolizumab ^3,4^. However, only 40% of ATC patients harbor this mutation^1,5^, leaving 60% of patients without effective systemic therapy. Those with stage IVB non-BRAF mutated disease are usually considered for surgery when feasible (rare) and/or chemoradiation^6^ +/-adjuvant pembrolizumab^7^. Unfortunately, 50% of ATC patients present with stage IVC (metastatic) disease and therefore there is an unmet need in these patients.

It has become increasingly clear that single agent therapies should not be utilized in patients with metastatic ATC^8^. Lenvatinib, a potent VEGFR inhibitor that is approved for differentiated thyroid cancer in the United States (US), has been studied in a clinical trial for ATC in the US. Single agent lenvatinib failed to show efficacy with a response rate of 2.9% and a median OS of 3 months^9^. Other single agent kinase inhibitors have shown similar, disappointing results^10-12^. The immune checkpoint inhibitor, spartalizumab, was tested in ATC. The response rate was 19% and median OS was 6 months. Those with higher PDL1 score had improved outcomes^13^. Since most ATC tumors express PDL1, it seems logical to combine immune checkpoint inhibitors with other agents. In a small, retrospective series of 6 ATC patients treated with lenvatinib + pembrolizumab, 4 partial responses (PR) were reported^14^. A larger prospective clinical trial was performed in Germany (ATLEP) and was presented at ESMO in 2022. This trial showed an impressive response rate of 52% and median overall survival of 11 months^15^.

We designed a single center, phase 2 trial in the US with Lenvatinib + pembrolizumab in patients with non-BRAF mutated ATC to determine the efficacy and safety of this combination.

## Methods

This was an open-label, single center, phase 2 clinical trial with lenvatinib + pembrolizumab 400mg every 6 weeks. The lenvatinib recommended starting dose was 20 mg daily, however patients who are considered to be at high risk of bleeding, per the treating physician’s discretion, could start at a reduced dose of lenvatinib 10 mg po daily and later dose escalate to 14 mg and then 20 mg po daily if deemed medically necessary and safe to do so. Patients were allowed to dissolve lenvatinib in water and swallow or place in a feeding tube if they were not able to swallow whole capsules. Up to 18 cycles of pembrolizumab were allowed on trial but lenvatinib was permitted to continue. Eligibility criteria include non-BRAF^V600E^ mutated ATC with metastatic disease, although patients with locoregional only disease were eligible if they were unwilling to undergo surgery or radiation. RECIST measurable disease was not necessary for trial entry (but active disease was), given that ATC patients are rare and the primary endpoint did not include the response rate. Patients were required to have an Eastern Cooperative Oncology Group (ECOG) performance status of <2 and normal organ function. We excluded patients with uncontrolled hypertension, significant cardiovascular impairments, clinically significant hemoptysis or tumor bleeding within 2 weeks, open wounds/fistulas, previous anti-angiogenic targeted therapy, untreated brain metastases and active autoimmune disease that required systemic treatment in the past 2 years. Patients were allowed to undergo surgical resection of the primary tumor during the trial if deemed feasible. Lenvatinib was held 1 week prior to the surgery and restarted 2 weeks after surgery, once adequate wound healing had occurred.

The primary efficacy endpoint was overall survival (OS). Secondary endpoints included progression-free survival (PFS), confirmed objective response rate (ORR) by investigator assessment per RECIST v1.1, and safety. Restaging scans were performed at 6 weeks from trial entry and then every 12 weeks. Responses were confirmed at ≥4 weeks. Treatment past progression was permitted if the patient was clinically benefiting.

The statistics were based on a comparison to the historical median OS of 3 months in ATC patients (based on the phase 2 single agent lenvatinib trial results^9^) with a goal to improve OS by 3 months (to 6 months median OS) using lenvatinib + pembrolizumab combination treatment. Assuming the accrual rate of one patient per month and 12 months additional follow-up after the last patient was enrolled, with 25 patients we had 90% power using 1-sided 5% alpha for a one-sample log rank test, and the number of events required of 18 (calculated using ST-plan). OS was defined as defined as the time from the start of the treatment to the date of death. Participants without documented death at the time of the final analysis were censored at the date of the last follow-up. PFS was defined as the time from the start of the treatment to the first occurrence of disease progression or death from any cause, whichever occurs first, as determined by an independent radiologist according to RECIST v1.1. Kaplan-Meier method was used to estimate the survival function. We estimated the median survival time and its 95% confidence intervals (CI). Adverse events (AEs) were assessed as defined by Common Terminology Criteria for Adverse Events (CTCAE), version 5.0. We performed the futility monitoring of OS using a Bayesian method^16^. That is, the trial would stop early if given the interim data, the probability of the median OS being longer than the historical control by 3 months is less than 4%, i.e., Pr(mE > mS + 3 | data) < 0.04.

The protocol was approved by the MD Anderson Cancer Center institutional review board and was conducted in accordance with the principles of the Declaration of Helsinki and Good Clinical Practice Guidelines. All patients provided written informed consent prior to study enrollment.

## Results

From 12/16/2021 to 2/3/2025, a total of 44 patients were screened for the clinical trial of which 25 were enrolled. Reasons for screen failures were: 14 did not meet eligibility criteria and 5 decided to pursue lenvatinib + pembrolizumab outside of the trial. The baseline patient characteristics are listed in Table 1. The median age was 62 years and 64% of patients were male. Seventy-two percent of patients had stage IVC disease at the time of their diagnosis, while 4% and 24% had stage IVA and IVB, respectively; however, all patients had distant metastatic disease at the time of study entry. Since the primary tumor is the initial cause of death in most patients with ATC, nearly all patients (96%) received treatment to the disease in the neck (either surgery or radiation) prior to study entry in order to avoid an airway complication. This included 17/25 (60%) who received palliative radiation (14-30 Gy), 3/25 (12%) who received full dose radiation (60-66 Gy), and 11/25 (44%) who had upfront surgical resection of the primary tumor. Two patients had brain metastases at baseline and received radiation to the brain prior to trial entry. The median number of cycles completed on the trial was 6 (range: 1-25).

**Table 1.**
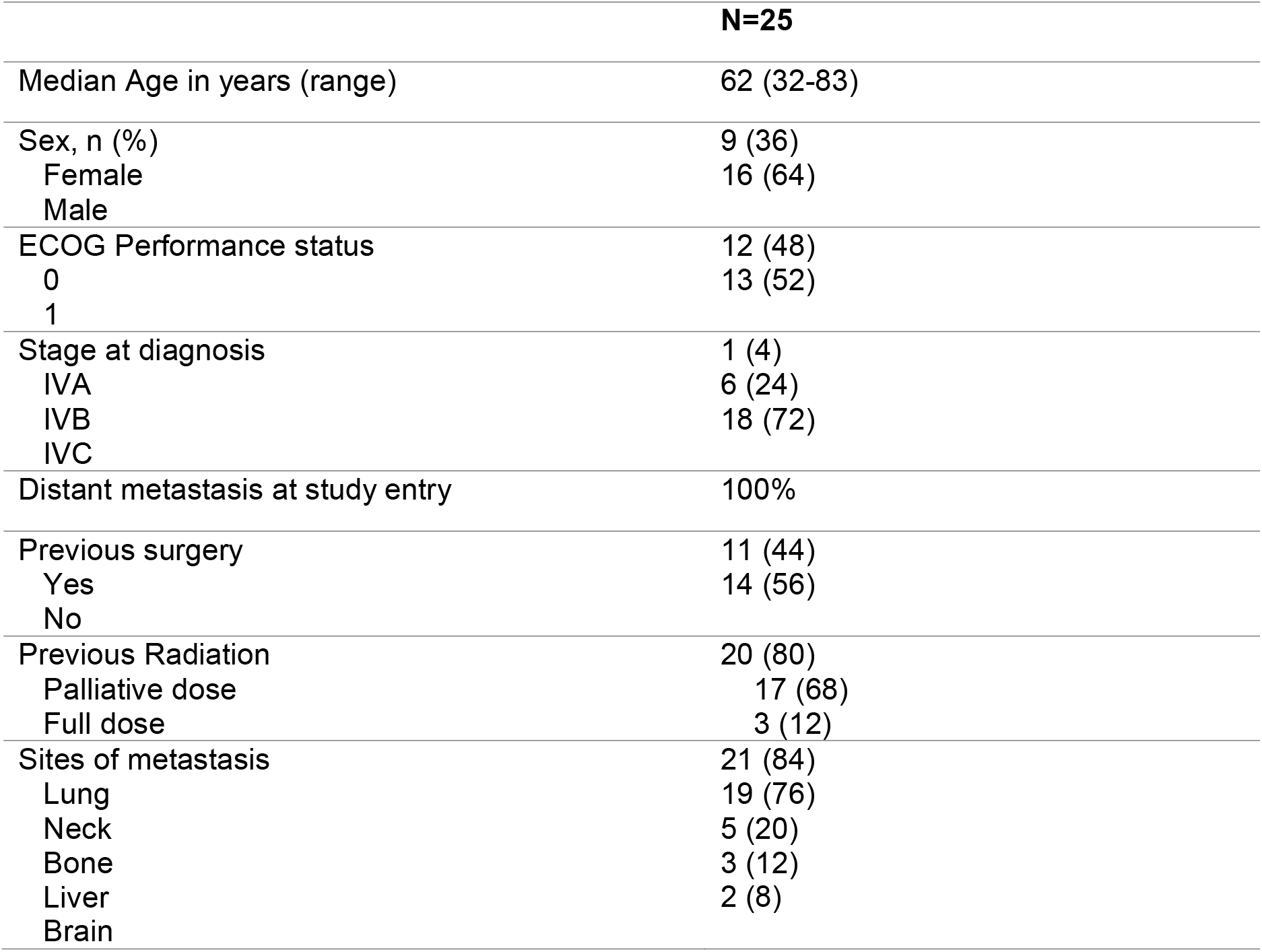
Patient Demographics.

### Efficacy

As of the cut-off date of 11/4/2025, 15 deaths and 19 patients with PD were observed. The median follow-up time for patients still alive was 17.5 months (range: 8.8 to 41.4 months). The median OS was 11.4 months (95% CI: 7.8 to 35.6; figure 1A). Notably, the lower bound of the 95% confidence interval (CI) exceeds the historical median OS of 3 months, suggesting that the one-sided p-value is <0.025. At 1 and 2 years, 50% and 31% were alive. The median PFS was estimated to be 5.4 (95%CI: 3.8, 11.0; figure 1B) months.

**Figure 1.**
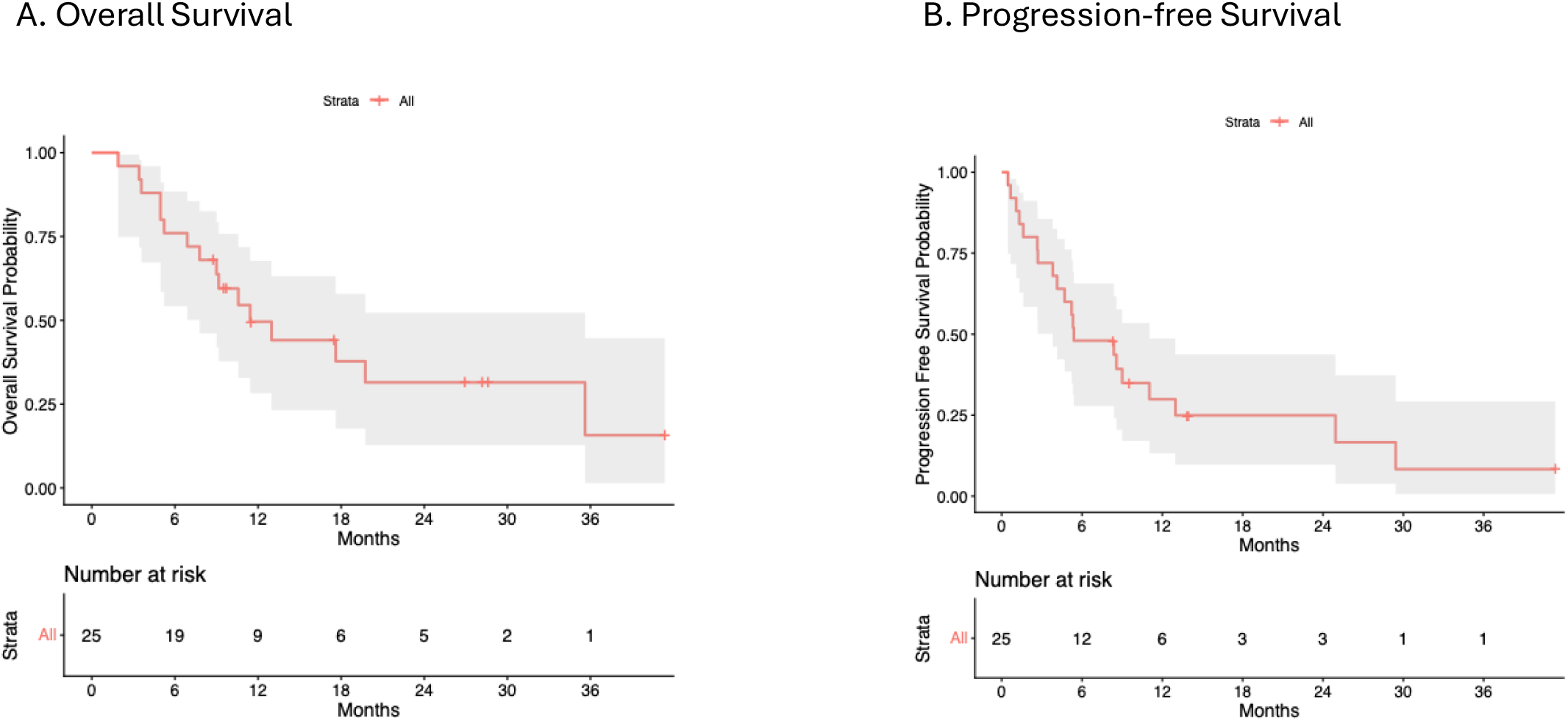
Kaplan-Meier plots showing the median overall survival and progression-free survival with lenvatinib + pembrolizumab in non-BRAF mutated ATC patients.

The best overall response rate was 36% (9/25), including 1 complete response and 8 partial responses (figure 2). Another 36% (9/25) had stable disease as the best response and there were 7 (24%) patients whose best response was progression. The trial protocol allowed patients to start at a reduced dose of lenvatinib per the discretion of the treating physician and this occurred in the majority of patients (68%).

**Figure 2.**
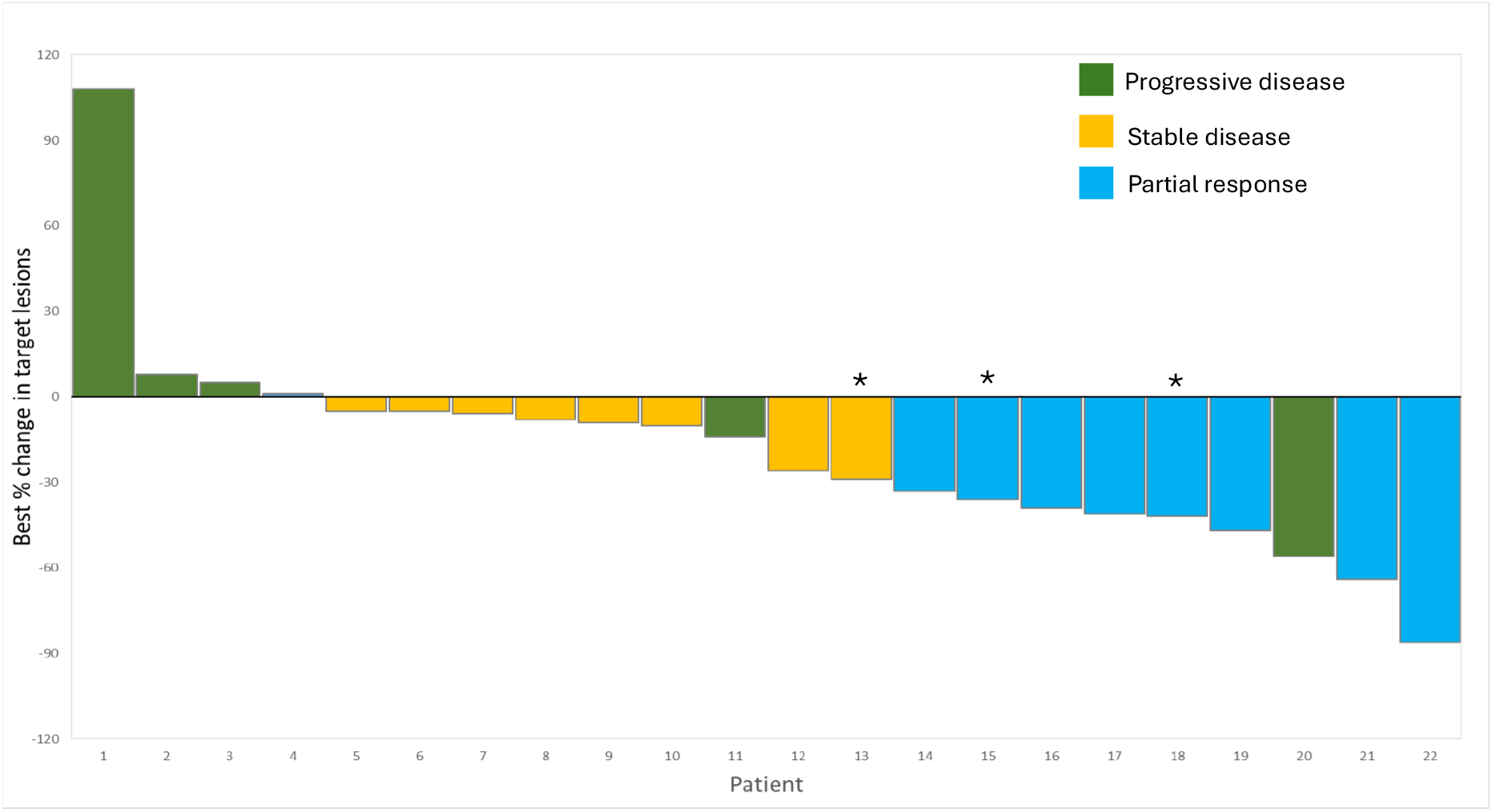
Waterfall plot showing the best overall response in target lesions in 23 patients were evaluable for response. Two patients had non-target lesions only—one with complete disappearance of non-target lesions and the other with stable disease in non-target lesions. One patient died early. Four patients had sufficient response in the primary thyroid tumor to facilitate surgical resection (asterisk; the 4th patient had non-measurable primary tumor and is not represented on the plot).

Of the 17 patients who underwent palliative radiation to the neck prior to study entry, 75% (12/17) had regression or stable disease in the neck while on clinical trial, including 3/17 (18%) patients with sufficient tumor shrinkage to allow for resection of the primary tumor (lobectomy). There was no viable tumor on surgical pathology in 2 of 3 of these patients’ tumor specimens (one with PTEN mutation and one with NRAS Q61R mutation). Another patient with a PTEN mutation had residual ATC with extensive treatment effect as well as PTC in the surgical specimen. One additional patient (HRAS Q61R mutated) who never received palliative radiation was able to undergo surgery during the clinical trial. This patient had no ATC in the specimen but a 0.2 cm papillary thyroid cancer (PTC) was identified. All 4 patients who underwent surgical resection of primary tumor remain alive at the time of data cut-off. Figure 3 shows the pre-treatment and pre-surgical images for the 4 patients.

**Figure 3.**
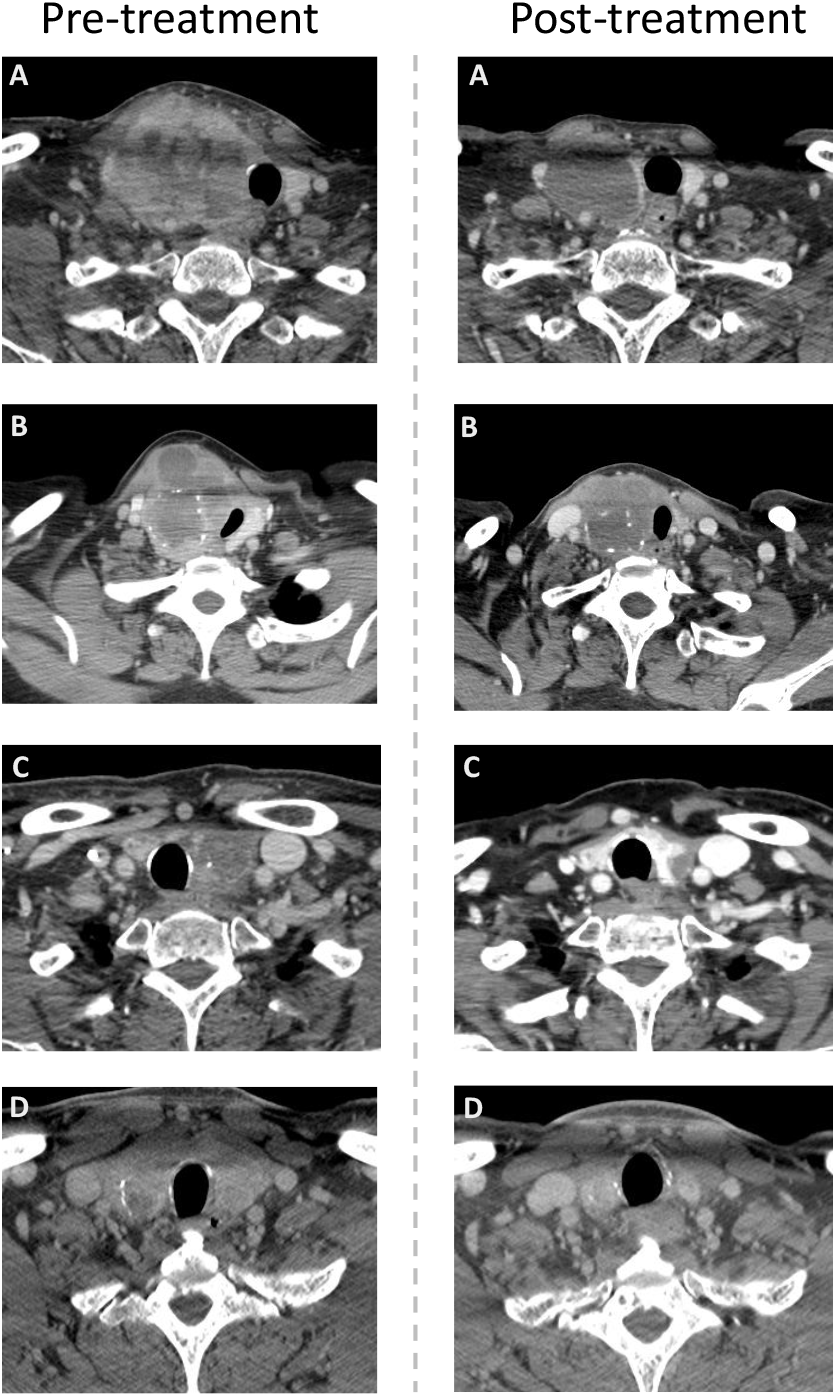
Four patients who had sufficient tumor shrinkage to allow for surgical resection of the primary tumor. Paired pre- and post-treatment contrast enhanced axial CT images of four patients (A-D) obtained at baseline and prior to resection. Pre-treatment images demonstrate large tumor volume at baseline with mass effect and extension to critical structures. Post-treatment images demonstrate significant reduction in tumor volume with decreased mass effect allowing for safe resection. The heterogenous nodular right thyroid tumor in patient A with encasement of the common carotid artery which was unresecteable at baseline is substantially improved on the post treatment scan with predominantly non-enhancing necrotic tumor remaining, allowing for safe resection. Similarly, there is decreased tumor volume with improved invasion of the sternocleidomastoid and subcutaneous soft tissues seen in patient B on the post treatment study. Infiltrative tumor extending into the left tracheoesophageal groove is improved on the post treatment scan for patient C with the tumor more well-defined and confined to the thyroid, allowing for safe resection. Treatment response is also seen in patient D with diffusely decreased tumor volume seen as diffusely decreased size of the thyroid gland after treatment. All patients had lobectomy and 3 of 4 patients (A, B, and C) had a complete ATC pathologic response to therapy.

### Biomarkers

The most common driver mutation was in RAS (48%), followed by PTEN (16%) and oncofusions (12%). There was no identifiable driver mutation in 20% of the patients. PDL1 was performed in 23/25 patient tumor specimens, of which 87% had a PDL1 score of ≥1% (range 5-100%). Of the 3 patients who had no PDL1 expression in tumor, 2 had progression as their best response and 1 had a partial response. The median tumor mutation burden (TMB) was 2 (range 0-36).

### Safety

All patients experienced at least one AE while on clinical trial. The most common AEs were weight loss (52%), hypertension (48%), anorexia and palmar-plantar erythrodysesthesia (each 36%) and proteinuria (32%). The most common grade ≥ 3 AE was hypertension, occurring in 20% of patients.

There were 2 serious bleeding events (1 upper GI and 1 tracheal hemorrhage), 2 GI perforations, and 2 fistulas. One patient had acute kidney injury with hypertension and proteinuria which was attributed to pembrolizumab. The patient recovered and continued on lenvatinib. Other notable SAEs were: pneumatosis intestinalis (grade 3, n=1), cholecystitis (grade 3, n=1), sepsis (grade 3, n=1), pulmonary embolism (grade 3, n=1), and tracheal mucositis (grade 3, n=1).

## Discussion

The present prospective clinical trial shows efficacy of lenvatinib + pembrolizumab combination therapy in non-BRAF mutated ATC, showing a median OS of 11 months compared to the previously published phase 2 clinical trial, which demonstrated 3 months OS with lenvatinib alone^9^. The combination led to a 36% ORR, including one complete response.

The ATLEP trial using lenvatinib + pembrolizumab in ATC had a similar median OS however the ORR was higher (52%). As this study remains unpublished, it is not clear if these were confirmed partial responses. There were also no patients where progression was the best response—an unusual finding in this aggressive patient population. In the ATLEP trial, the starting dose of lenvatinib was 20mg, whereas in our trial, most patients started at the 10mg dose level. This may have led to the difference in response rates, however, the median OS was not different. It is also unclear what the driver mutations were in the ATLEP trial. In our trial, 48% of patients had RAS mutations, and differences in patient populations could have accounted for the differences in ORR between ATLEP and our trial. Patients with RAS mutations have the worst prognosis when compared to BRAFV600E mutated or non-BRAF/non-RAS mutated ATC (adjusted HR 2.64). Specifically, patients with RAS-mutated ATC tumors have a 1-year survival of 31%, compared to 64% in BRAF mutated^1^. Another series of ATC patients with RAS mutation showed that 70% of these patients present with distant metastatic disease, and of those who do not, they develop distant metastases at a median time of <6 months^17^. The very high rate of distant metastatic disease, coupled with the lack of effective targeted therapies against RAS mutations, likely accounts for why these patients have a shorter median OS.

In our trial, 80% of patients had some form of radiation to the neck, while 44% had previous surgery. Due to local complications from rapid tumor growth, it is our standard of practice to treat patients with either upfront surgery (when resectable with limited distant disease) or palliative radiation for locoregionally threatening disease in the neck. Thus, if patients with distant metastatic non-BRAF mutated ATC present without previous surgery or radiation, we typically recommend upfront palliative radiation (14-30 Gy) to the primary tumor. Tan et al^18^, reported a case series of 5 patients who received pembrolizumab every 3-4 weeks, followed by palliative radiation “QUAD-shot” (14 Gy over 4 fractions), which was repeated every 3-4 weeks for a total of up to 56 Gy. This resulted in 2 CRs and 2 PRs and long-term survival in 2 patients.

In our study, patients received palliative RT followed by lenvatinib + pembrolizumab, resulting in 75% locoregional control. Of the 4 patients in the trial who had surgery after lenvatinib + pembrolizumab, 3 had no viable ATC on the surgical pathology and all 4 patients are alive. Multiple series have shown a survival advantage to surgical resection in ATC, independent of stage^19-21^. In BRAF mutated ATC patients, neoadjuvant dabrafenib/trametinib +/-pembrolizumab followed by surgery has been shown to improve OS^4,22^. Thus, we believe that palliative radiation followed by lenvatinib + pembrolizumab should be studied as a strategy, not only to facilitate rapid locoregional control, but also as a neoadjuvant therapy in non-BRAF mutated ATC.

Unfortunately, bleeding events and fistulas were seen in a minority of patients despite lowering the dose of lenvatinib. These events are eventually fatal because lenvatinib must be stopped in these situations, and this often leads to progression. Other adverse events were as expected for this combination. In patients who already have bleeding or are at high risk of bleeding or fistula due to the location of the tumor, an alternative systemic therapy regimen may be considered. In the clinical trial with ipilimumab + nivolumab by Sehgal et al, no fistulas were reported^23^. Although the ATC cohort was small, this immunotherapy combination appeared to be effective.

In conclusion, the combination of lenvatinib plus pembrolizumab shows efficacy in patients with metastatic, non-BRAF mutated ATC compared to lenvatinib alone. The addition of palliative dose radiation (14-30 Gy) prior to lenvatinib + pembrolizumab results in excellent locoregional control and should be a standard of care for this patient population.

## Data Availability

All data produced in the present study are available upon reasonable request to the authors

## Acknowledgments

We wish to thank the patients and their supportive friends and families, as well as the research staff who helped care for our patients.

Funding for this trial was provided by Merck Investigator Studies Program (MISP), Gateway for Cancer Research, and the Petrick-MD Anderson Cancer Center Philanthropic Funds.

## Conflicts of Interest Declaration

MEC: Research funding from Genentech, Exelixis, Regerneron, Merck; Advisory board: Bayer, Exelixis, Novartis, Fore. Consulting for Thryv.

RD: Research funding from Eisai, Merck, Exelixis. Consulting for Novartis and Bristol Myers Squibb.

MZ: Research funding from Merck, Lilly, and Exelixis.

SH: Speaker for Lily; Research funding Exelixis and Regeneron.

AM: Research funding from Jazz Pharmaceuticals, THRYV Therapeutics, NABORS Industries

RF: personal fees from Regeneron, Sanofi, Merck Serono, Elevar Therapeutics, Prelude Therapeutics, Eisai Inc, Remix Therapeutics, and Coherus BioSciences; nonfinancial support from Ayala Pharmaceuticals, EMD Serono, ISA, Genentech/Roche, Merck Serono, Pfizer, Viracta, and Gilead outside the submitted work.

NLB: Eisai – advisory board; research funding (through subcontract); Novartis – research funding through subcontract; Lilly – speaker funds; Exelixis – advisory board.

MGM, MIH, GBG, JRW, PI, SL, VB, MS, MN: No COI to declare

## Notes

### Competing Interest Statement

MEC: Research funding from Genentech, Exelixis, Merck; Advisory board: Bayer, Exelixis, Novartis, Fore. Consulting for Thryv.
RF: personal fees from Regeneron, Sanofi, Merck Serono, Elevar Therapeutics, Prelude Therapeutics, Eisai Inc, Remix Therapeutics, and Coherus BioSciences; nonfinancial support from Ayala Pharmaceuticals, EMD Serono, ISA, Genentech/Roche, Merck Serono, Pfizer, Viracta, and Gilead outside the submitted work.
NLB: Eisai: advisory board; research funding (through subcontract); Novartis: research funding through subcontract; Lilly: speaker funds; Exelixis: advisory board.
MDW: Bayer: research support.
MGM, JRW, PI, SL, VB, MS: No COI to declare

### Clinical Trial

NCT04171622

### Author Declarations

The University of Texas MD Anderson Cancer Center IRB gave ethical approval for this work.

